# Frequency and Longitudinal Course of Autonomic Reflex Testing Abnormalities in Isolated REM Sleep Behavior Disorder

**DOI:** 10.1101/2024.11.19.24317581

**Authors:** Hash Brown Taha, Jennifer Zitser, Mitchell G. Miglis

## Abstract

**Study Objectives:** Autonomic dysfunction is common across the α-synucleinopathies including isolated RBD (iRBD), however the presence, severity and distribution of autonomic dysfunction as a risk factor for phenoconversion in iRBD remains unclear. We aimed to characterize autonomic reflex testing (ART) abnormalities in a cohort of participants with iRBD and assess their potential as biomarkers for predicting phenoconversion risk.

**Methods:** We performed ART on 45 individuals with iRBD and evaluated the ability of individual ART components (sympathetic cholinergic, cardiovagal, sympathetic adrenergic) to predict phenoconversion using univariate and multivariate predictive models, both alone and combined with measures of olfaction, cognition, motor function, and skin biopsy assessment of dermal synuclein.

**Results:** Forty-one individuals with iRBD were enrolled (age 66.7 ± 7.4 yrs, 27% female), and followed annually for an average of 2.9 ± 2.4 yrs, with four participants lost to follow-up. Eight participants with iRBD phenoconverted during their follow-up period (3 Parkinson’s disease, 4 dementia with Lewy bodies and 1 multiple system atrophy), yielding a phenoconversion rate of 6.6% per year. Eighty seven percent of iRBD participants had an abnormal baseline ART, and 100% had an abnormal follow-up ART. A combination of MDS-UPDRS III score and cardiovagal dysfunction (abnormal heart rate variability with deep breathing) best predicted phenoconversion (AUC = 0.77, 95% CI: 0.59–0.94).

**Conclusions:** ANS dysfunction was common and spanned all domains of autonomic function. Baseline cardiovagal dysfunction was most affected and predictive of phenoconversion, especially if combined with motor examination. Longitudinal studies with larger sample sizes are needed to confirm these findings.

## Introduction

The α-synucleinopathies are a group of neurodegenerative diseases characterized by the accumulation of α-synuclein in the central and peripheral nervous system. They include Parkinson’s disease (PD), dementia with Lewy bodies (DLB), multiple system atrophy (MSA), and pure autonomic failure (PAF). (1) REM sleep behavior disorder (RBD) is common in the α-synucleinopathies, (2, 3) and isolated RBD (iRBD), or RBD in those not meeting criteria for PD, DLB, or MSA, often precedes the motor manifestations of these disorders by up to several decades, thus presenting the ideal population in which to deploy disease-modifying therapies. (4-6) A recent meta-analysis estimated the risk of phenoconversion of iRBD to a neurodegenerative disorder to be 33.5%, 82.4% and 96.6% at 5, 10.5 and 12 years follow-up, respectively. (7) (8) However, despite recent advances in biomarkers, (9-14) phenoconversion rates to PD, DLB or MSA can vary substantially among those with iRBD, making prognosis challenging for clinicians.

Autonomic nervous system (ANS) dysfunction is another common prodromal feature of the α-synucleinopathies, (15, 16) and is also highly prevalent in those with iRBD. (17) Indeed, several studies have reported high co-occurrence rates of iRBD and ANS dysfunction. (18, 19) Despite this, very few studies have evaluated objective, standardized autonomic reflex testing (ART) in individuals with iRBD. (3, 20) In this study, we aimed to address this knowledge gap by evaluating cross-sectional rates as well as longitudinal progression of ANS dysfunction in a well-phenotyped cohort of video polysomnogram (vPSG)-confirmed individuals with iRBD using ART.

## Methods

Participants with iRBD were prospectively recruited and consecutively enrolled from the Stanford Sleep Disorders Clinic. All participants were clinically evaluated for dream enacting behaviors. The diagnosis of iRBD was based on ICSD-3 criteria (21) with confirmation of REM sleep without atonia (RSWA) on attended video polysomnography (vPSG) with a combination of submentalis, upper extremity, and lower extremity electromyography (EMG). RSWA was evaluated according to SINBAR criteria, (22, 23) using a 50% cut-off for loss of REM sleep atonia using a combination of chin and upper extremity EMG, according to International Classification of Sleep Disorders-3 scoring criteria. All patients with a 4% apnea-hypopnea index (AHI) of >15/h were excluded unless obstructive sleep apnea was adequately treated with positive airway pressure therapy. Exclusion criteria included diagnoses of PD, DLB, MSA, dementia, autonomic failure (pure autonomic failure idiopathic orthostatic hypotension), or conditions associated with autonomic failure including uncontrolled diabetes, vitamin B12 deficiency, or alcohol abuse. RBD onset was defined as onset of dream enacting behavior, recalled by the participants and/or their bedpartners.

Participants were evaluated annually to assess phenoconversion and all underwent a comprehensive neurological examination including the Movement Disorders Society Unified Parkinson’s Disease Rating Scale (MDS-UPDRS) part III and the Montreal Cognitive Assessment (MoCA) at baseline and at follow up visits. Olfaction was quantified with the University of Pennsylvania Smell Identification Test (UPSIT)-40 at baseline only. Due to the COVID-19 pandemic, some follow-up visits were conducted remotely during this time period. All participants had in-person follow-up at the time of their most recent visit. Most participants (68.2%) also underwent skin biopsy to assess dermal phosphorylated α-synuclein (p-syn), as previously described. (24)

Participants underwent a standard battery of ART, including measures of sympathetic cholinergic function (quantitative sudomotor axon reflex testing (QSART) using a Q-Sweat machine (WR Medical Electronics, Stillwater, MN)), cardiovagal function (heart rate variability with deep breathing (HRVdb)), Valsalva ratio (VR), and sympathetic adrenergic function (Valsalva blood pressure (BP) analyses, and 10-minute head-up tilt (HUT) table testing at an angle of 70-degrees, with continuous blood pressure (BP) and heart rate (HR) monitoring with finger plethysmography confirmed with an automated cuff sphygmomanometer over the brachial artery). The HUT ΔHR/ΔSBP ratio was calculated by dividing the change in HR by the change in systolic BP (SBP) at 3 minutes, a validated measure of neurogenic orthostatic hypotension (nOH), as previously described. (25) Medications known to affect ART were held for at least five half-lives prior to testing. Orthostatic hypotension (OH) was defined as a ≥20 mm Hg decrease in SBP and/or ≥10 mm Hg decrease in diastolic BP (DBP) sustained across two consecutive minutes of HUT. Classic OH was defined as a sustained BP drop within 3 of HUT, and delayed OH as a sustained blood pressure drop after 3 min of HUT. Supine hypertension (sHTN) was defined as a SBP ≥140 mm Hg and/or DBP ≥90 mm Hg), as per consensus criteria. (26, 27)

The composite autonomic severity scale (CASS), a well-defined quantification of autonomic function testing results comprised of cardiovagal, adrenergic, and sudomotor domains, (28) was calculated for each patient as described previously. (24) CASS is used to evaluate the severity of autonomic dysfunction by scoring these three key domains separately: cardiovagal (0-3 points), adrenergic (0-4 points), and sudomotor (0-3 points). The total score ranges from 0 to 10 and allows for a graded assessment of autonomic impairment. A score of 0 indicates normal autonomic function, while higher scores reflect varying degrees of autonomic failure, with scores closer to 10 indicating severe autonomic dysfunction. A subset of participants agreed to repeat ART to evaluate longitudinal progression of ANS dysfunction.

Phenoconversion to PD, DLB or MSA was defined according to standard criteria and made by the participant’s treating neurologist. For 7 phenoconverters (3 PD, 4 DLB) this was by the investigators (MM, JZ), however one participant was diagnosed by their outside treating movement disorders specialists (1 MSA).

Statistical significance for continuous variables was evaluated using an independent t-test or the Mann-Whitney U-test, depending on the normality of data distribution, and Fisher’s exact test was used for categorical variables. Effect size was calculated using Cohen’s d. (29) Receiver operating characteristics (ROC) analyses were conducted using a binomial logistic regression without least absolute shrinkage and selection operator (LASSO) for univariate models and with LASSO variable selection for multivariate models (MDS-UPDRSIII, QSART volume at the forearm, proximal and distal leg, and foot, HRVdb, VR, baseline SBP, HUT ΔSBP 3 min, HUT ΔSBP/HR 3 min, HUT ΔHR/SBP 3 min, Valsalva BP baseline to phase 2 BP minimum, Valsalva BP phase 2 recovery, Valsalva phase 4 BP overshoot, CASS sudomotor, cardiovagal, sympathetic and total score). (30) The sensitivity and specificity were calculated at the optimal cutoff that maximizes Youden’s index. (31) Prediction error was compared using the Akaike information criterion (AIC) for the ROC models; a lower AIC indicates higher predictive accuracy, resulting in a more reliable model, with differences around -2 to -3 indicating better models. (32) All analyses and graphs were conducted in RStudio (version 2022.12.0+353).

## Results

Four participants were lost to follow-up during the study period, and we were not able to determine their phenoconversion status. Forty-one individuals with iRBD were enrolled (age 66.7 ± 7.4 yrs, 27% female, disease duration 6.9 ± 5.6 yrs), and followed annually for an average of 2.9 ± 2.4 yrs. Eight iRBD participants phenoconverted during their follow-up period (3 PD, 4 DLB and 1 MSA), yielding a phenoconversion rate of 19.5% (6.6% per year). Both phenoconverters and non-phenoconverters were similar in age, gender, body mass index, age of disease onset, disease duration, antidepressant usage, dermal p-syn positivity, cognitive function (MoCA) or hyposmia (UPSIT-40). Participants who phenoconverted had significantly higher motor (MDS-UPDRSIII) dysfunction. MoCA scores < 26 were seen in 6/33 (18%) of non-phenoconverters and 1/8 (12.5%) of phenoconverters, rates of MCI in iRBD congruent with prior studies. (33) These results are summarized in Table 1.

**Table 1.**
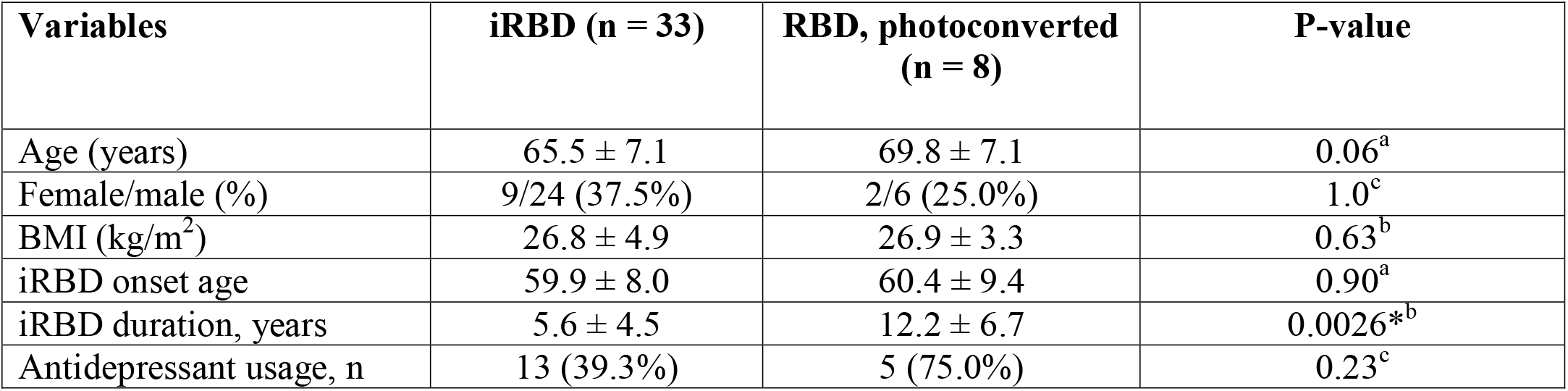
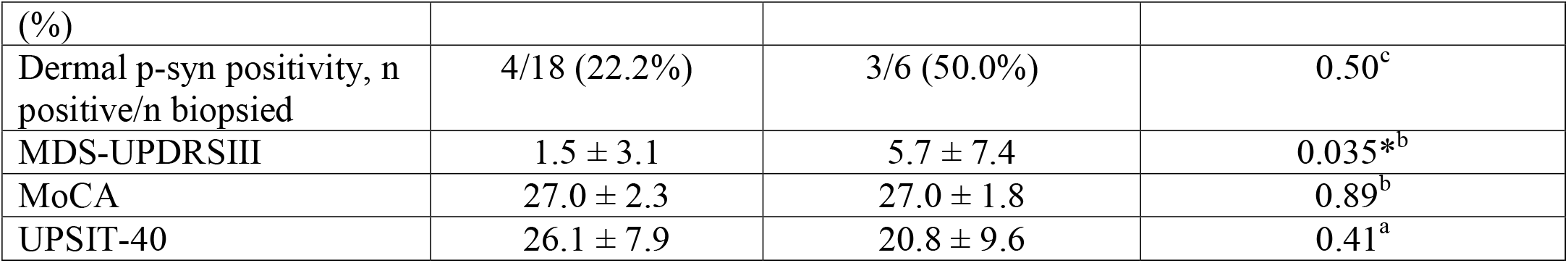
Demographic and clinical feature comparisons between individuals with isolated RBD (iRBD) and those who have phenoconverted to a synucleinopathy (RBD). Statistical tests were run based on the data’s normality. ^a^ Independent t-test (normally distributed data). ^b^ Mann-Whitney U-test (non-normally distributed data). ^c^ Fisher’s exact test. Statistical significance (*) is shown for p-value < 0.05. One participant declined sudomotor testing, while cardiovagal function could not be calculated in one participant due to paced cardiac rhythm. BMI = body mass index, p-syn = phosphorylated α -synuclein, MDS-UPDRSIII = Movement Disorders Society-Unified Parkinson’s Disease Rating Scale III, MoCA = Montreal Cognitive Assessment, UPSIT-40 = University of Pennsylvania Smell Identification Test-40.

In our cohort (including the 4 lost to follow-up) ART was abnormal in 39/45 (87%). Sympathetic cholinergic (sudomotor) testing was abnormal in 15/44 (34%), with 40% of those exhibiting a length-dependent pattern and 60% exhibiting a non-length dependent pattern of sweat loss. Cardiovagal function (HRVdb and/or VR) was abnormal in 25/44 (57%). Sympathetic adrenergic function was abnormal in 24/45 (53%), with 22/45 (49%) demonstrating an excessive SBP fall in phase II and 23/45 (51%) demonstrated lack of SBP overshoot in phase IV of the Valsalva maneuver. Orthostatic hypotension (OH) was seen in 12/45 (27%) on HUT, with 10 exhibiting classic OH (sustained blood pressure drop within 3 min of HUT) and three delayed OH (sustained blood pressure drop after 3 min of HUT). Using ΔHR/SBP <0.5 at minute 3 of HUT as a marker of nOH, 6/45 participants (13%), or 50% of those with OH, met criteria for nOH. Using OH + lack of phase IV Valsalva BP overshoot as a marker of nOH, 8/45 (18%) met criteria for nOH. Only 3 participants (7%) had lack of phase IV Valsalva BP overshoot on testing + OH + ΔHR/SBP <0.5. Only HRVdb was significantly higher in phenoconverters. These results are summarized in Table 2.

**Table 2.** Autonomic reflex testing results between individuals with isolated RBD (iRBD) and those who have phenoconverted to a synucleinopathy (RBD). Statistical tests were run based on the data’s normality. ^a^ Independent t-test (normally distributed data). ^b^ Mann-Whitney U-test (non-normally distributed data). ^c^ Fisher’s exact test. Statistical significance (*) is shown for p-value < 0.05. One participant declined sudomotor testing, while cardiovagal function could not be calculated in one participant due to paced cardiac rhythm. QSART = quantitative sudomotor axon reflect testing, HRVdb= heart rate variability with deep breathing, P2 = phase 2 Valsalva maneuver, P4 = phase 4 Valsalva maneuver, HUT = head-up tilt, SBP = systolic blood pressure, CASS = Composite Autonomic Severity Score.

| Variables | iRBD (n = 39) | RBD, photoconverted (n = 6) | P-value (Cohen's d effect size) |
| --- | --- | --- | --- |
| QSART volume forearm | 1.3 ± 0.75 | 1.4 ± 0.59 | 0.48<sup>a</sup> (-0.24) |
| QSART volume proximal leg | 0.71 ± 0.35 | 0.51 ± 0.26 | 0.10<sup>a</sup> (0.58) |
| QSART volume distal leg | 0.59 ± 0.41 | 0.61 ± 0.36 | 0.72<sup>b</sup> (-0.061) |
| QSART volume foot | 0.54 ± 0.43 | 0.46 ± 0.28 | 0.73<sup>b</sup> (0.47) |
| HRVdb (bpm) | 9.7 ± 5.5 | 6.9 ± 2.4 | 0.036<sup>*a</sup> (0.56) |
| Valsalva ratio | 1.5 ± 0.29 | 1.4 ± 0.20 | 0.82<sup>b</sup> (0.18) |
| Valsava baseline to P2 minimum (mmHg) | 37.0 ± 22.1 | 42.5 ± 22.2 | 0.59<sup>a</sup> (0.25) |
| Valsava P2 recovery (mmHg) | 5.3 ± 11.9 | 9.5 ± 14.0 | 0.97<sup>b</sup> (-0.34) |
| Valsava P4 overshoot (mmHg) | 7.4 ± 9.3 | 5.2 ± 8.5 | 0.56<sup>b</sup> (0.24) |
| HUT baseline SBP (mmHg) | 139.0 ± 12.8 | 145.1 ± 13.5 | 0.28<sup>a</sup> (-0.47) |
| HUT baseline HR (bpm) | 63.7 ± 10.9 | 65.4 ± 12.3 | 0.89<sup>b</sup> (-0.14) |
| HUT ΔSBP 3 minutes (mmHg) | 14.6 ± 10.9 | 16.0 ± 11.5 | 0.76<sup>b</sup> (-0.12) |
| HUT ΔHR 3 minutes (bpm) | 10.5 ± 7.2 | 15.1 ± 14.5 | 0.49<sup>A</sup> (-0.51) |
| Ratio HUT ΔHR/ΔSBP 3 minutes | 1.4 ± 2.0 | 1.8 ± 3.0 | 0.78<sup>b</sup> (-0.15) |
| CASS sudomotor | 0.90 ± 1.2 | 0.62 ± 1.1 | 0.71<sup>b</sup> (0.23) |
| CASS cardiovagal | 1.5 ± 1.2 | 1.4 ± 1.1 | 0.85<sup>b</sup> (0.075) |
| CASS sympathetic | 1.5 ± 1.1 | 1.5 ± 1.1 | 0.93<sup>b</sup> (0.030) |
| CASS total | 3.9 ± 2.6 | 3.7 ± 2.5 | 0.86<sup>a</sup> (0.071) |

Six iRBD participants agreed to undergo repeat ART an average of 4.6 ± 1.2 yrs after initial ART testing, all of whom retained their iRBD phenotype. All participants had abnormal ART results (33% sympathetic cholinergic, 100% cardiovagal and 83% sympathetic adrenergic impairment; Figure 1), worsening in all domains. OH was seen in 50% (two classic and one delayed, also present on participants’ prior baseline HUT). One individual with a normal HUT developed delayed OH on follow-up testing.

**Figure 1.**
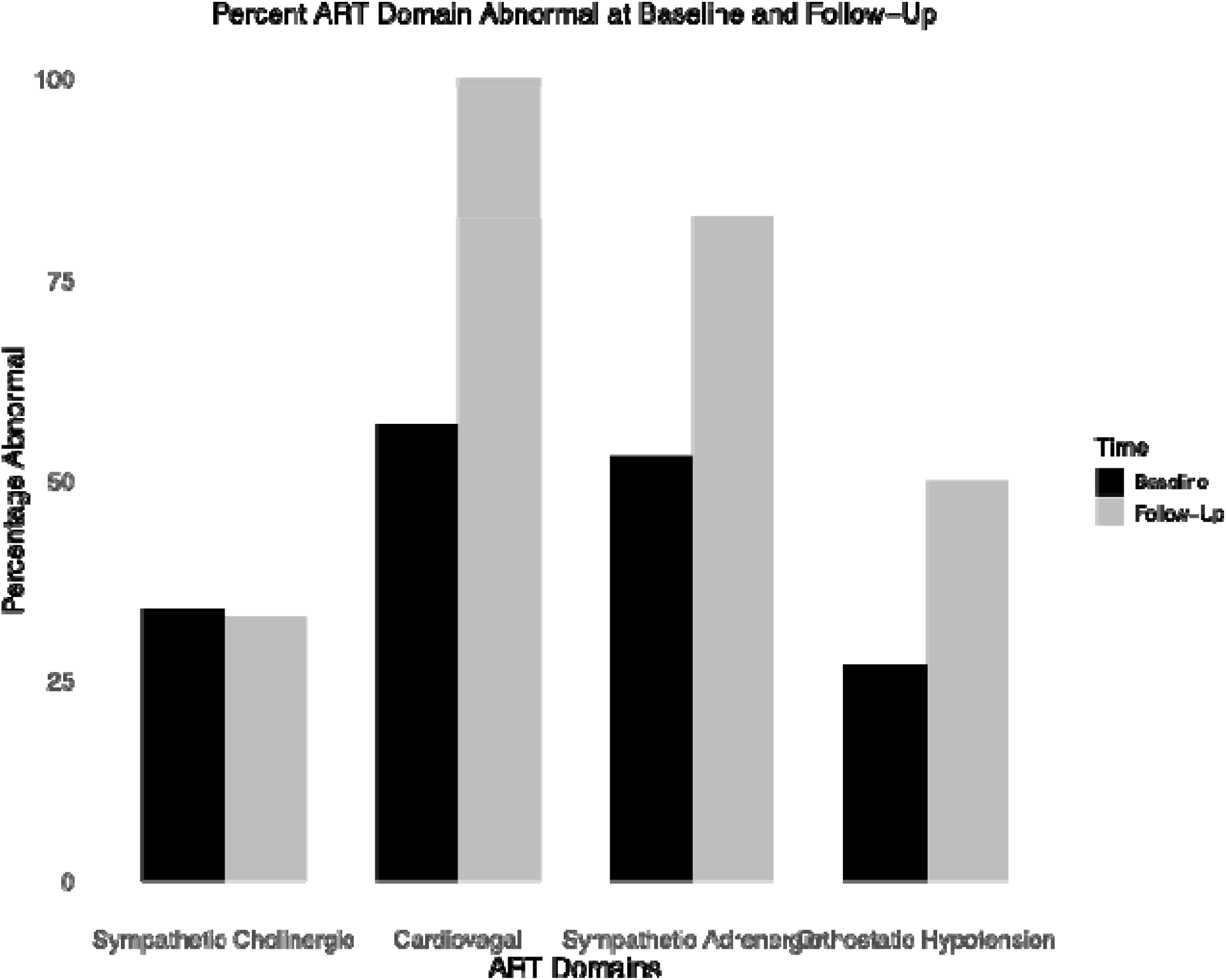
Percentage of autonomic reflex testing abnormality at baseline and on follow-up.

We performed univariate and multivariate ROC models to evaluate the ability of each biomarker to differentiate between phenoconverters and non-phenoconverters (Table 3). Baseline MDS-UPDRSIII total score provided fair predictive accuracy (AUC = 0.728), while biomarkers of sympathetic cholinergic dysfunction (QSART volume at the forearm, proximal and distal leg, and foot) mostly provided poor predictive accuracy except for QSART proximal leg volume (AUC = 0.664). Of all cardiovagal dysfunction assessments, HRVdb was the strongest predictor of phenoconversion (AUC = 0.667). Variables evaluating sympathetic adrenergic, orthostatic hypotension and those from the CASS examination provided the poorest predictive accuracy.

**Table 3.**
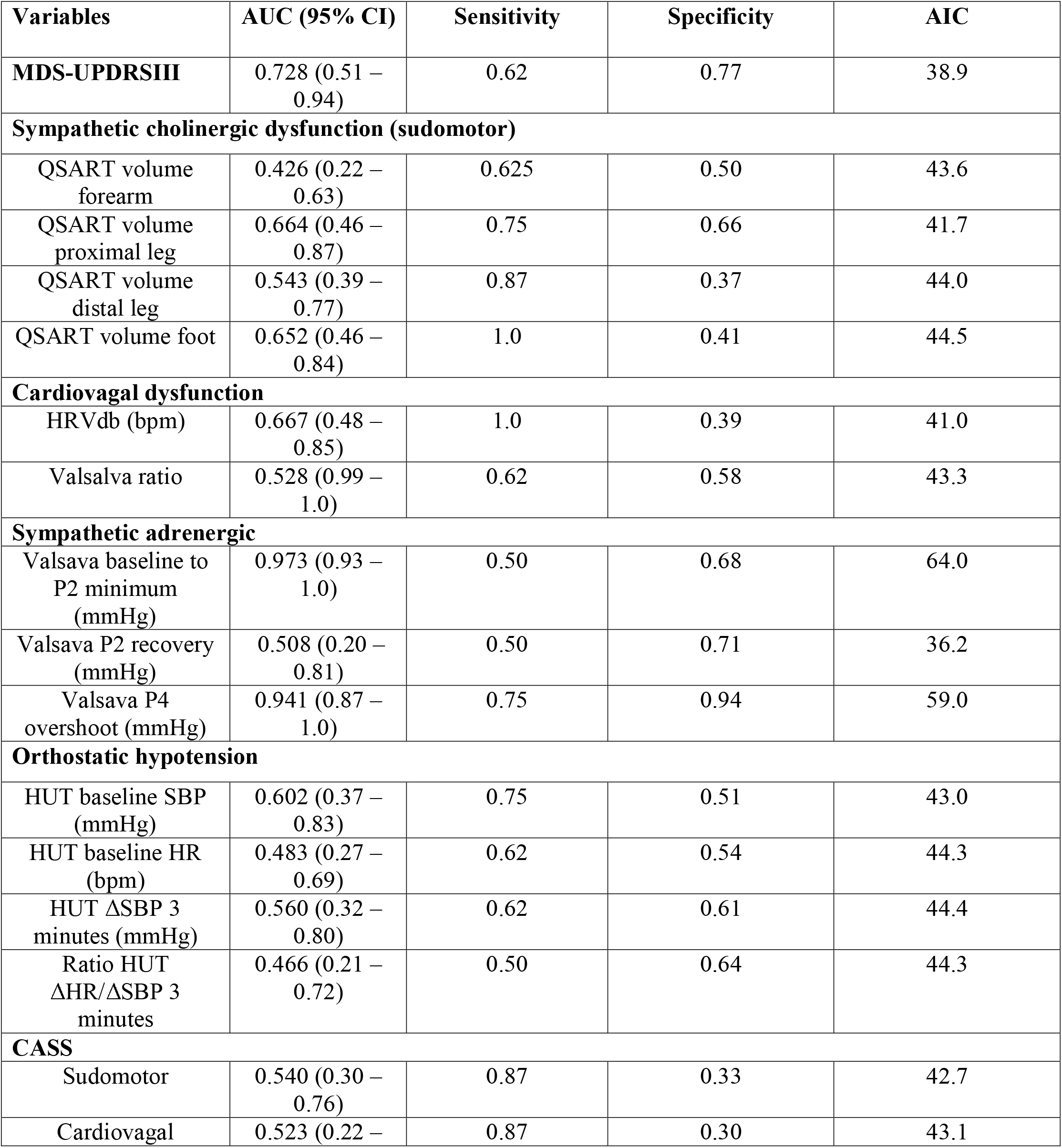
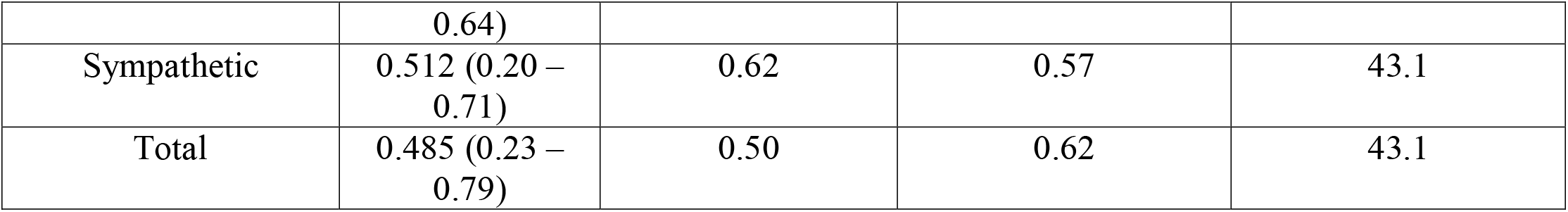
Univariate receiver operating characteristics models. AUC – area under the curve. AIC – Akaike information criterion. MDS-UPDRSIII = Movement Disorders Society-Unified Parkinson’s Disease Rating Scale III, QSART = quantitative sudomotor axon reflect testing, HRVdb= heart rate variability with deep breathing, P2 = phase 2 Valsalva maneuver, P4 = phase 4 Valsalva maneuver, HUT = head-up tilt, SBP = systolic blood pressure, HR = heart rate, CASS = Composite Autonomic Severity Score.

To evaluate which combination of motor and ART abnormality best predicted risk of phenoconversion with ROC analyses using a binomial logistic regression and LASSO variable selection, MDS-UPDRSIII and HRVdb were selected as the best predictors of iRBD to α-synucleinopathy phenoconversion (AUC = 0.77, 95% CI: 95% CI: 0.59–0.94; Figure 2). These results did not change to a significant degree after excluding the single individual who phenoconverted to MSA.

**Figure 2.**
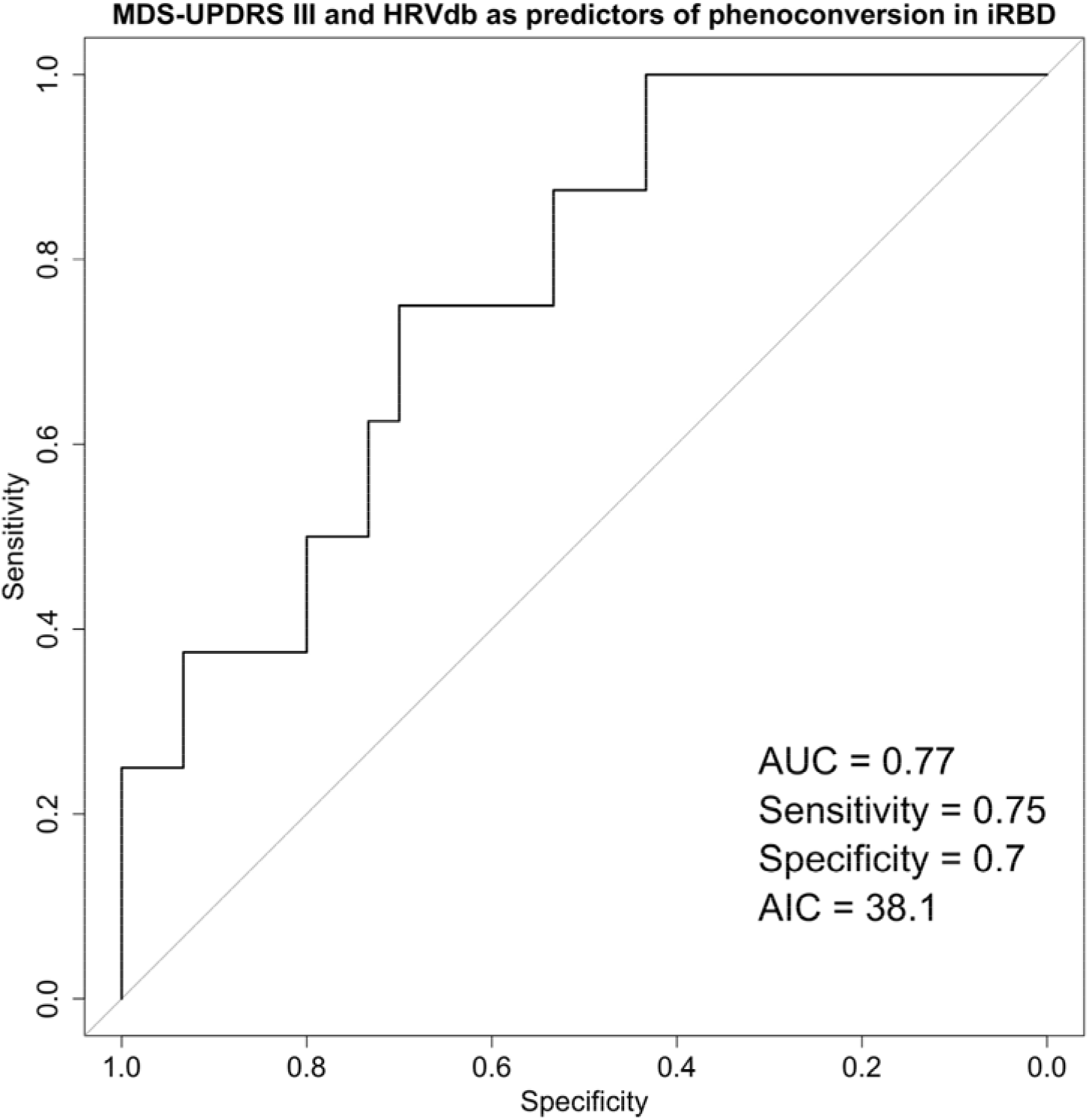
Receiver operating characteristics (ROC) analysis using a binomial logistic regression with LASSO variable selection. The model selected MDS-UPDRSIII and HRVdb and as the best predictors for phenoconversion in iRBD. AUC – area under the curve. AIC – Akaike information criterion. MDS-UPDRSIII = Movement Disorders Society Unified Parkinson’s Disease Rating Scale Part III. QSART = quantitative sudomotor axon reflex testing. HRVdb = heart rate variability with deep breathing.

## Discussion

In this study, we followed 45 participants with iRBD for an average of 2.9 ± 2.4 years, during which 8 participants phenoconverted (3 PD, 4 DLB, 1 MSA). ANS dysfunction was common and spanned all domains of autonomic function. In addition, our phenoconversion rate (19.5% overall and 6.6% per year) was similar to other studies, which note a rate of 6-8%. (33, 34) While other groups have also reported ANS dysfunction in iRBD using measures of HR variability, (35, 36) cardiac scintigraphy, (37) orthostatic stand testing, (17) and ART, (15) to our knowledge our report details ART on the largest cohort of iRBD participants to date.

Sudomotor responses, a marker of peripheral sympathetic cholinergic function, was abnormal in 34% of our cohort, with most exhibiting a non-length dependent pattern. Similar findings of non-length dependent sudomotor abnormalities have been seen in other small ART cohorts, (38, 39) suggesting that synuclein-driven neuronal degeneration occurs not only in a non-contiguous fashion in iRBD, but in both central and peripheral structures early in the disease process.

Cardiovagal function was abnormal in over half of our cohort and was the most affected domain of autonomic function (Figure 1). These findings are consistent with other ART studies in iRBD, one of which found cardiovagal abnormalities in 14/17 (82%) of their cohort. (39) Parasympathetic dysfunction is known to occur early in PD and DLB and constipation is one of the earliest prodromal features, present in up to 95% of those with iRBD. (33) Impaired cardiovagal function may also be an early prodromal feature, and likely progresses with disease severity. Indeed, on follow up ART testing, 100% of iRBD participants in our cohort had cardiovagal impairment, compared to 57% at baseline. Cardiovagal abnormalities were also the only domain that predicted phenoconversion in our cohort. One retrospective study found that more severe cardiovagal (assessed using CASS) impairment on ART predicted higher risk of phenoconversion to DLB than PD, (38) however we did not find this to be the case in our cohort, perhaps limited by a ceiling affect given most of our cohort exhibited cardiovagal abnormalities at baseline.

We found high rates of OH (27%) on HUT, identical to rates of OH seen in the North American Prodromal Synucleinopathy (NAPS) cohort of 340 iRBD participants based on a three-minute active stand test (27%). (17) In this study, we used the Δ HR/SBP at 3 min of active standing to estimate nOH, finding that 21% met this criterion, accounting for the vast majority of those with OH. However, while demonstrating high sensitivity and specificity of predicting nOH in a cohort of participants with autonomic failure, (25) ΔHR/SBP has not been validated across other cohorts, including in those with iRBD, as ART is not performed in the NAPS cohort.

A blunted phase IV BP overshoot on Valsalva maneuver is a validated marker of sympathetic adrenergic impairment, and combined with OH on HUT is generally accepted as the gold standard for the diagnoses nOH. Using this combination, 18% of our iRBD participants met criteria for nOH, suggesting that ΔHR/SBP alone may overestimate nOH in patients with iRBD. However, sympathetic impairment was quite common in our cohort, with 51% demonstrating a blunted phase IV response. Like cardiovagal impairment, sympathetic impairment may present as a continuum along the disease spectrum of iRBD, first manifesting as an abnormal Valsalva BP response, followed eventually by delayed OH, then classic OH as the disease progresses towards phenoconversion.

While follow-up ART was limited to only six participants, we found persistent and severe ANS dysfunction on repeat testing (Figure 1), with one individual developing delayed OH on repeat HUT, suggesting ANS dysfunction in iRBD is progressive and may serve as a biomarker of disease progression. In support of this, one longitudinal study (40) found that 44% of those with delayed OH eventually develop classic OH over a follow-up period of ten years, again emphasizing the concept of disease progression from prodromal to manifest disease states of autonomic dysfunction in the synucleinopathies.

Limitations of our study include the fact that our sample was a single-site cohort with limited longitudinal follow up period, limited sample size of those who underwent follow-up ART, and lack of repeat ART at regular intervals in all participants. Despite these limitations, our study further highlights the strong bidirectional link between iRBD and autonomic failure. Furthermore, ANS dysfunction is common in iRBD and may be used to inform phenoconversion risk. Specifically, a combination of motor (MDS-UPDRS III total score) and cardiovagal (HRVdb) dysfunction may serve as prognostic biomarkers in iRBD. Future studies are needed to confirm these findings in larger longitudinal cohorts.

## Data Availability

The data that support the findings of this study are available on request from the corresponding author. The data are not publicly available due to privacy or ethical restrictions.

## Acknowledgments

HBT: study concept, drafting and editing of manuscript, statistical analysis; JZ: editing of manuscript; MGM: study concept, drafting and editing of manuscript.

